# Random Forest Model for Predicting Post-Lockdown Antenatal Depression Risk: A Cross-Sectional Study of Pregnant Women in China

**DOI:** 10.64898/2026.05.23.26353929

**Authors:** Yijin Pan, Hai Lin, Takae Hirono, Yifan Yang, Yang Liu, Yinqiang Zhang

**Author notes:** Corresponding author: Yinqiang Zhang, No.1 Shuaifuyuan, Dongcheng District, Beijing 100730, China.

## Abstract

**Background:** As lockdown measures was eased, pregnant women faced an elevated risk of COVID-19 infection, potentially impacting their mental health. This study aimed to investigate the prevalence of antenatal depression (AD) post-lockdown and develop predictive models for AD risk using machine learning.

**Methods:** A cross-sectional study utilizing the Edinburgh Postnatal Depression Scale was conducted in Beijing and Guizhou, China, from January to August 2023. Data was randomly split into training and test datasets (6:4 ratio), with logistic regression (LR), Support Vector Machine (SVM), K-Nearest Neighbors (KNN), Random Forest (RF), eXtreme Gradient Boosting (XGBoost), and Gradient Boosting Decision Tree (GBDT) models trained and compared. The best model underwent further examination, including SHapley Additive exPlanations (SHAP) for feature importance, calibration curve (CC) for discrimination, and decision curve analysis (DCA) for clinical benefit.

**Results:** The effective response rate was 91.07% (459/504), with 25.7% (118/459) testing positive for AD. Multivariate analysis identified “sleep disorders,” “family support level,” and “COVID-19 symptom severity” as independent predictors. RF model showed the highest area under the curve in both training (0.842) and testing (0.724) datasets, with SHAP emphasizing the greatest impact of “sleep disorders” on AD. The RF model’s calibration (*P* > 0.05) and clinical utility across thresholds (8%-95% and 10%-58%) were confirmed by CC and DCA, respectively.

**Conclusions:** AD strongly correlated with “sleep disorders,” “family support level,” and “COVID-19 symptom severity” post-lockdown, and the EPDS-based RF model effectively predicted AD risk.

## 1. Introduction

According to the “2022 World Mental Health Report” of World Health Organization (WHO), the coronavirus disease 2019 (COVID-19) pandemic has led to an escalation in the global burden of mental disorders, with cases of severe depression rising by 28%. In response to this pandemic, China implemented various prevention and control measures, including enforcing home quarantine, implementing flexible work arrangements, and promoting telecommuting [1]. Although these measures significantly helped alleviate depression caused by the unknown virus during the early stages of the epidemic, inconveniences such as restricted mobility, reduced social interactions, and information overload disrupted individual’s daily routines. Amid ongoing stress and uncertainty, the mental health challenges stemming from the epidemic persist, particularly affecting pregnant women [2]. Pregnancy represents a unique period for women of reproductive age, during which they entail significant identity and role adjustments. If negative emotions during pregnancy are not promptly identified and appropriately managed, they can develop into antenatal depression (AD), which is primarily characterized by persistent low mood and loss of interest. Untreated AD can pose serious risks to maternal and fetal health, increasing the likelihood of pregnancy complications and adverse pregnancy outcomes [3–4]. In severe cases, AD can lead to suicidal behavior in pregnant women, which is a major cause of direct maternal mortality during pregnancy, thus straining the healthcare system. Additionally, antidepressants such as selective serotonin reuptake inhibitors may potentially affect fetal central nervous system development via the placenta. The efficacy and safety of these medications remain controversial among pregnant women [5]. Given the high risks associated with AD and the limitations of treatment methods, the WHO recommends early identification and intervention for pregnant women predisposed to AD.

Before the COVID-19 pandemic, the reported prevalence of AD abroad was approximately 9.9% [6], while it typically ranged from 12.5% to 18.9% in China [7–8]. The combined effects of the pandemic and prenatal stress have exacerbated the psychological burden on pregnant women, leading to a significant increase in the incidence of AD during the early stages of the pandemic. For example, a meta-analysis that encompassed global surveys of negative emotions among pregnant women from December 2019 to February 2021 revealed a pooled prevalence of depression of 25.6%. This result was significantly higher than historical norms utilizing similar methodologies [2]. A study published recently found that women who were pregnant during the pandemic were nearly twice as likely to exhibit symptoms of depression compared to matched women who were pregnant prior to the pandemic. Factors at both the individual and community levels linked to socioeconomic disparities were associated with latent factors of COVID-19-related stress and adversity [9]. Another study conducted in the US indicated that peripartum women with pre-existing mental health diagnoses, as well as those experiencing pandemic-related health concerns or grief, were at increased risk for mental health symptoms [10]. Furthermore, logistic regression (LR) analysis conducted among pregnant women from Guangxi, China, indicated a mild depression rate of 27.4%, with pregnancy complications emerging as an independent risk factor while regular employment serving as a protective factor [11]. Another cross-sectional study conducted in Shaanxi, China, demonstrated a significant increase in hostility and depressive symptoms among peripartum women during the outbreak period, with a positive depression screening rate as high as 39.39% [12].

However, previous studies have mostly focused on the early stages of the pandemic outbreak, whereas relatively less attention given to the period following the easing of pandemic lockdown, and the modeling methods employed were often singular, lacking comparison of algorithm efficacy. By the end of 2022, pandemic-related restrictions were relaxed in China, and the focus of the country’s COVID-19 response shifted from “infection prevention” to “severe case prevention”. However, the easing of pandemic lockdown has increased the risk of pregnant women contracting COVID-19, which may lead to negative impacts on their psychological health. Based on this premise, the current study aims to utilize the Edinburgh Postnatal Depression Scale (EPDS) to investigate the prevalence of AD among pregnant women undergoing routine prenatal checkups in both Chaoyang District, Beijing, and Qiandongnan Prefecture, Guizhou Province, within approximately half a year following the easing of COVID-19 lockdown. Furthermore, the independent influencing factors of AD during this special period would be identified, and AD risk prediction models based on these factors would be established employing 6 machine learning (ML) algorithms, providing scientific evidence for the formulation of targeted intervention measures.

## 2. Material and methods

### 2.1 Study participants and sample size

This study employed a cross-sectional design, convenience sampling, and adhered to the principle of voluntary participation. Pregnant women receiving routine prenatal checkups at maternal and child health institutions in Chaoyang, Beijing, and Qiandongnan Prefecture, Guizhou Province, between January 20, 2023, and August 31, 2023, were eligible for participation in the questionnaire surveys. Inclusion criteria: (1) Confirmed pregnancy through clinical symptoms, laboratory, and imaging examinations; (2) Absence of reading, writing, or communication disorders, enabling independent completion of the questionnaire; (3) Acquisition of informed consent directly from the pregnant participants themselves. Exclusion criteria: (1) Previous diagnosis of depression or other mental disorders; (2) Presence of significant physical illnesses necessitating prolonged medication; (3) Refusal or termination of study participation. All participants in this study provided written informed consent prior to enrollment. The consent forms were developed in accordance with the Declaration of Helsinki guidelines and approved by the Institutional Review Board of Peking University (Approval Number: IRB00001052-22124).

Based on comprehensive reviews of existing research, the AD prevalence was set at 20%, with a confidence level of 95% and a permissible error of 0.02. Substituting these parameters into the sample size estimation formula of the cross-sectional study, with consideration of a 10% rate of questionnaire invalidity, the calculated minimum number of questionnaires is 426.

The cross-sectional study sample size estimation formula is as follows:

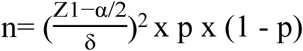

A total of 504 questionnaires were collected, resulting in 459 valid responses, representing an efficacy rate of 91.07%. This study has been approved by the Institutional Review Board of The survey questionnaire underwent multiple rounds of discussion by the research team and experts, and a pilot survey was conducted before the formal investigation to promptly refine the data collection form. Four investigators received homogeneous training and guidance to ensure familiarity with the content, methods, and precautions of data collection, thereby ensuring the accuracy of data collection. Two data managers are responsible for verifying and organizing the collected questionnaires, as well as supervising the quality control processing of raw data.

### 2.2 Risk and protective factors

A self-designed questionnaire encompasses three main aspects: (1) Sociodemographic characteristics, including age, ethnicity, pre-pregnancy occupation, current residence, type of housing, type of medical insurance, monthly household income, and marital status; (2) Pregnancy-related factors, comprising the intentionality of pregnancy, method of conception, delivery history (categorized into primiparous and multiparous women), presence of early pregnancy symptoms, presence of pregnancy complications, and behaviors such as smoking, drinking, and sleep quality. Sleep quality during pregnancy was assessed using the Pittsburgh Sleep Quality Index (PSQI), which was developed in 1989 [13]. The PSQI consists of 7 dimensions, each rated on a 4-point scale (0-3 points), resulting in a total score range of 0 to 21, where higher scores indicate poorer sleep quality. Following the recommendation from the investigation conducted on domestic pregnant women (Liu et al., 1996), a PSQI score of ≥7 was utilized as an indicator of sleep disorder among pregnant women in this study (Cronbach’s alpha = 0.75). Also assessed was the level of family support, which was defined as the degree of psychological and material support received from other family members during pregnancy, categorized into “high, middle, and low” levels according to a study conducted during the COVID-19 pandemic [14]. Other pregnancy-related factors included cohabitation status during pregnancy, primary decision-maker for antenatal care, proportion of pregnant women who received antenatal mental health education from professional institutions, and proportion of pregnant women who sought help from professional institutions for psychological issues. (3) COVID-19-related factors, encompassing 14 items covering the frequency and severity of infections (referring to authoritative guidelines both domestically and internationally, this study categorizes the severity of COVID-19 symptoms into three groups: “Not infected/Asymptomatic,” “Mild,” and “Moderate/Severe/Critical”), the impact due to COVID-19 (such as employment and economic effects, residential lockdown measures, and access to medical resources), concerns about COVID-19 (such as vertical transmission, threat to newborn health, and postpartum healthcare service needs), and duration of attention to COVID-19 news. These factors provide a comprehensive reflection of pregnant women’s attitudes and the degree of impact of the COVID-19 pandemic, serving as an important reference for the formulation of targeted support measures (Cronbach’s alpha = 0.83).

### 2.3 Outcomes

The EPDS was employed to evaluate the depressive status of pregnant women within the preceding week. Developed by Cox et al. in 1987, this scale comprises 10 items, each assessed on a 4-point scale (0-3 points), yielding a total score ranging from 0 to 30 points (Cox et al., 1987). A higher score correlates with a more serious level of depression. Previous studies have demonstrated its validity and reliability in detecting AD among pregnant women during the COVID-19 pandemic (King et al., 2023). Drawing from an early domestic study (Peng et al., 1994), this study adopted an EPDS score of ≥10 as a positive marker for AD screening.

### 2.4 Database establishment

A database was constructed using Microsoft Excel (Version 16.0.1), and initial screening was performed based on specific criteria as follows: (1) Each response was limited to one per social media platform, electronic device, and IP address; (2) Duplicate or excessively brief responses (duration < 90 seconds) were excluded; (3) Responses with birth dates beyond the 31st were omitted; (4) Variables with missing data proportions exceeding 5% in each column were deleted; (5) Illogical outlier values in the questionnaire were eliminated. The dataset was accessed for research purposes on May 10, 2024, with all analyses completed by June 10, 2024. Authors had access to identifiable participant information during the initial data collection phase, including IP addresses and device identifiers. All identifiers were retained in a secure, password-protected database separate from the primary research dataset. Identifiable information was only used for duplicate response detection and was permanently deleted after the initial screening process was completed. No identifiable participant information was accessible during data analysis or reporting stages.

### 2.5 Group comparison and variable selection

Uni- and multi-variate analyses were conducted using R software (Version 4.3.0). Pregnant women were categorized into AD (EPDS ≥ 10) and non-AD (EPDS < 10) groups based on EPDS scores. Differences in sociodemographic characteristics, pregnancy-related factors, COVID-19-related factor between groups were examined. Qualitative data were represented using frequency n (%) (due to rounding in composition ratios, the sum may not always equal 100%), comparison between groups employed chi-square tests or Fisher’s exact test. For quantitative data, normality tests using the Kolmogorov-Smirnov method were conducted first. Normally distributed data were expressed as X±S, and t-tests were used for group comparisons. Non-normally distributed data were presented as M (P25, P75), with comparisons conducted using Mann-Whitney U tests. Significant variables identified in univariate analysis (*P* <0.05) were incorporated into multivariate analysis to investigate their associations with AD, presented as odds ratios (OR) and 95% confidence intervals (CI).

### 2.6 Model establishment and evaluation

The model establishment and evaluation utilized the “sklearn” package in Python 3.10. The AD group (EPDS ≥10) and non-AD group (EPDS <10) were regarded as binary outcome variables. Variables deemed significant through multivariate analysis (*P* < 0.05), with acceptable levels of collinearity (VIF < 5), were considered independent predictive factors. The dataset was divided into a training dataset (N=276) and a test dataset (N=183) at a split ratio of 6:4. Six different artificial intelligence (AI) models were developed using the training dataset, including LR, Support Vector Machine (SVM), K-Nearest Neighbors (KNN), Random Forest (RF), eXtreme Gradient Boosting (XGBoost), and Gradient Boosting Decision Tree (GBDT). Performance of models were assessed using a range of indicators, including the receiver operating characteristic (ROC) area under the curve (AUC), precision, recall, accuracy, F1 score, sensitivity and specificity, with AUC value was employed to determine the optimal model. In the optimal model, the impact significance of each predictive factor on the outcome was assessed using SHapley Additive exPlanations (SHAP). Furthermore, the discriminative capacity of the optimal model was analyzed using a confusion matrix (CM) and calibration curve (CC), while the clinical utility of the model was evaluated via Decision Curve Analysis (DCA).

This study employed two-tailed tests, with statistical significance denoted by P < 0.05.

## 3. Results

### 3.1 Characteristics of the participants

Table 1 illustrates the sociodemographic characteristics of our survey respondents. Among the 459 pregnant women, the median age was 31.0 (28.0, 33.0) years old, with 66.4% were Han ethnicity. Furthermore, 93.2% reported having a pre-pregnancy occupation, 88.7% were presently residing in urban areas, 69.7% purchased their residence, 76.9% were covered by medical insurance for urban workers, and 59.0% had a total monthly family income ≥ 10,001 RMB.

**Table 1.**
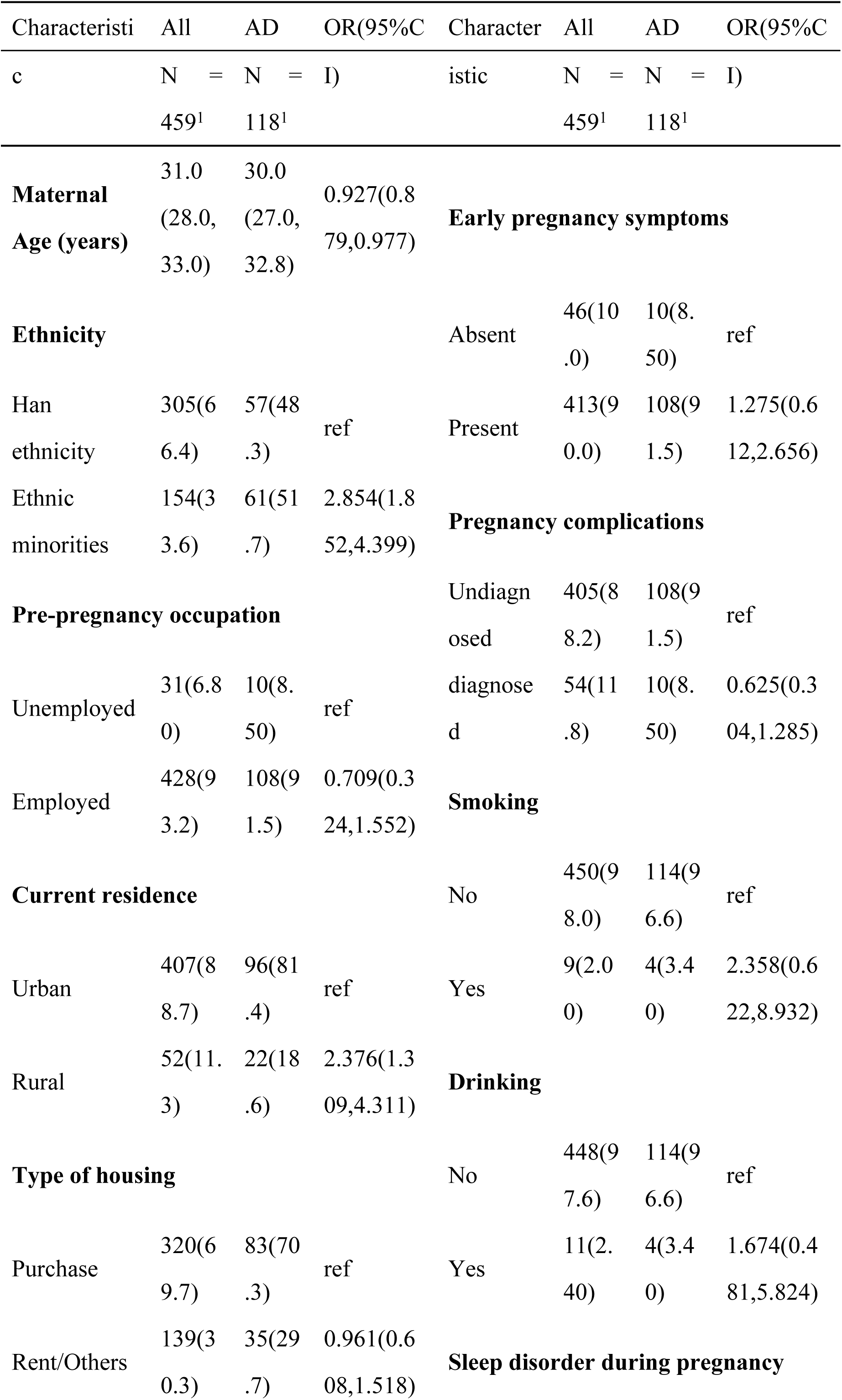
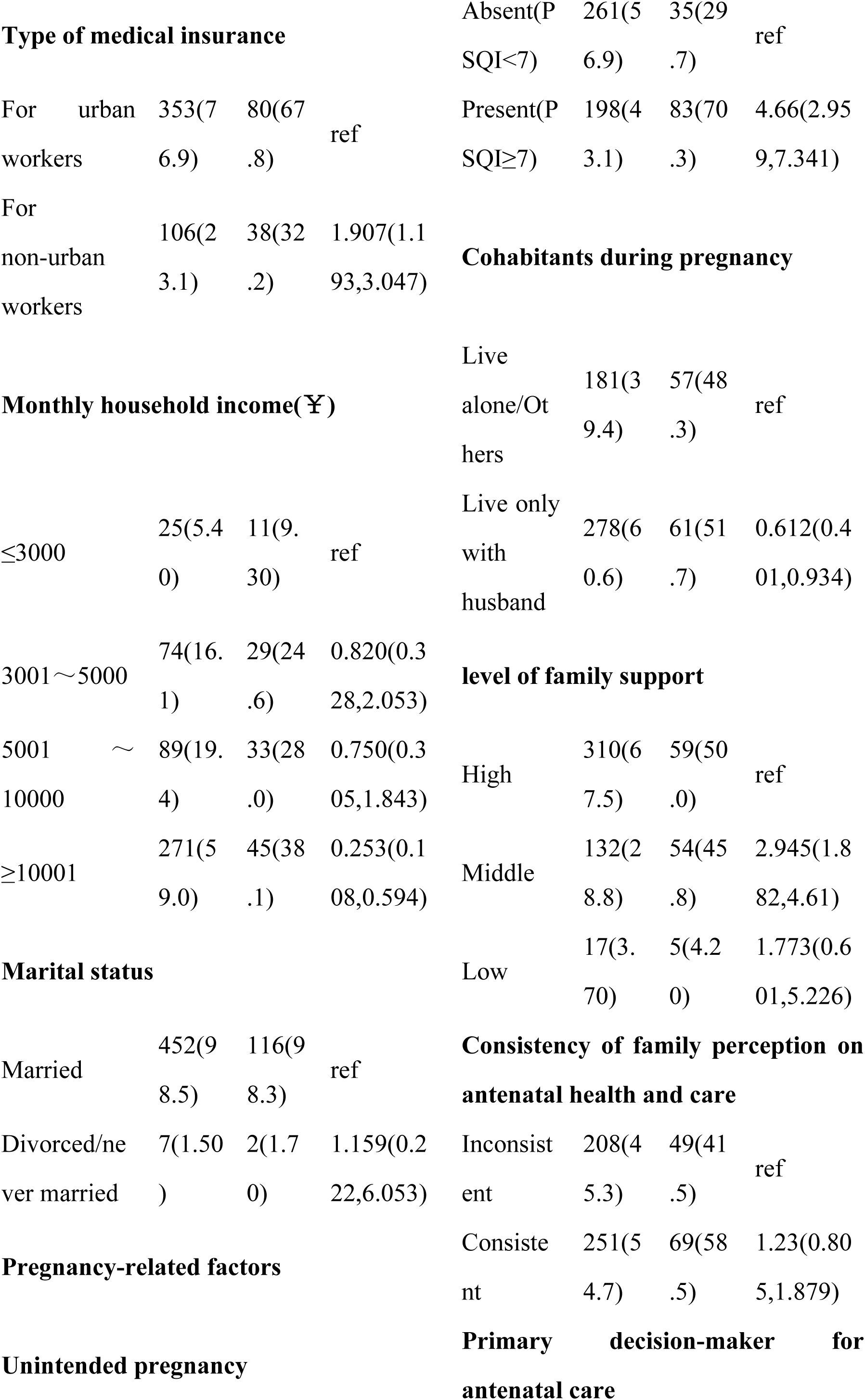
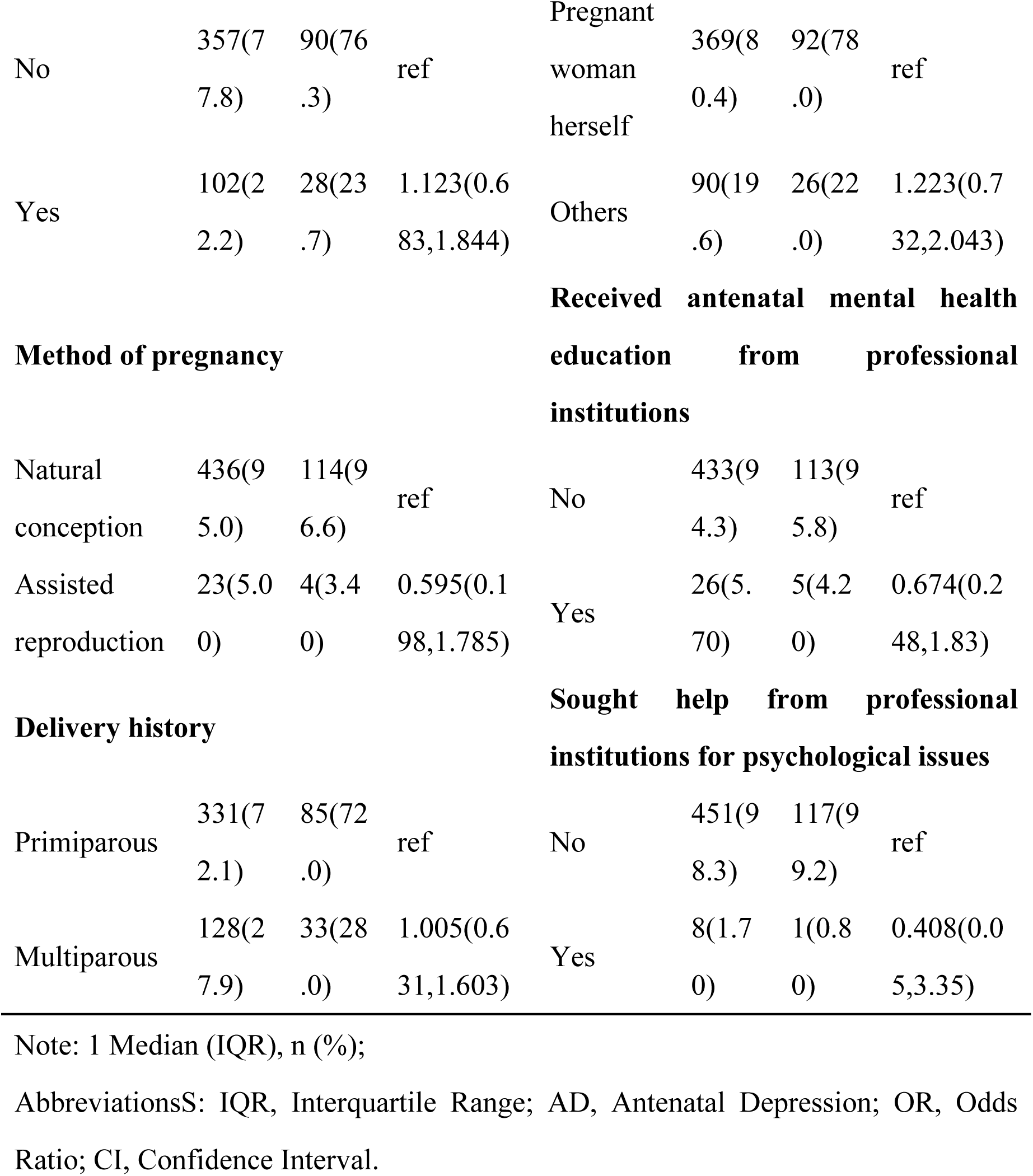
Basic characteristics in women with AD compared to women with Non-AD.

### 3.2 The prevalence of antenatal depression according to women’s characteristics

The study compared the prevalence of AD among women with distinct characteristics, encompassing sociodemographic characteristics (Table 1), pregnancy-related factors (Table 1), and COVID-19-related factors (Table 2). 25.7% (118/459) of pregnant women screened positive for AD. Comparing the AD group with the non-AD group, the former exhibited a younger median age, a higher proportion of ethnic minorities, a greater proportion residing in rural areas, more without medical insurance for urban workers, and a lower proportion with a total monthly family income ≥ 10,001 RMB. These differences were all statistically significant (*P* < 0.05). Conversely, no statistically significant differences were observed between the two groups concerning pre-pregnancy occupation, type of housing, and marital status (*P* > 0.05).

**Table 2.**
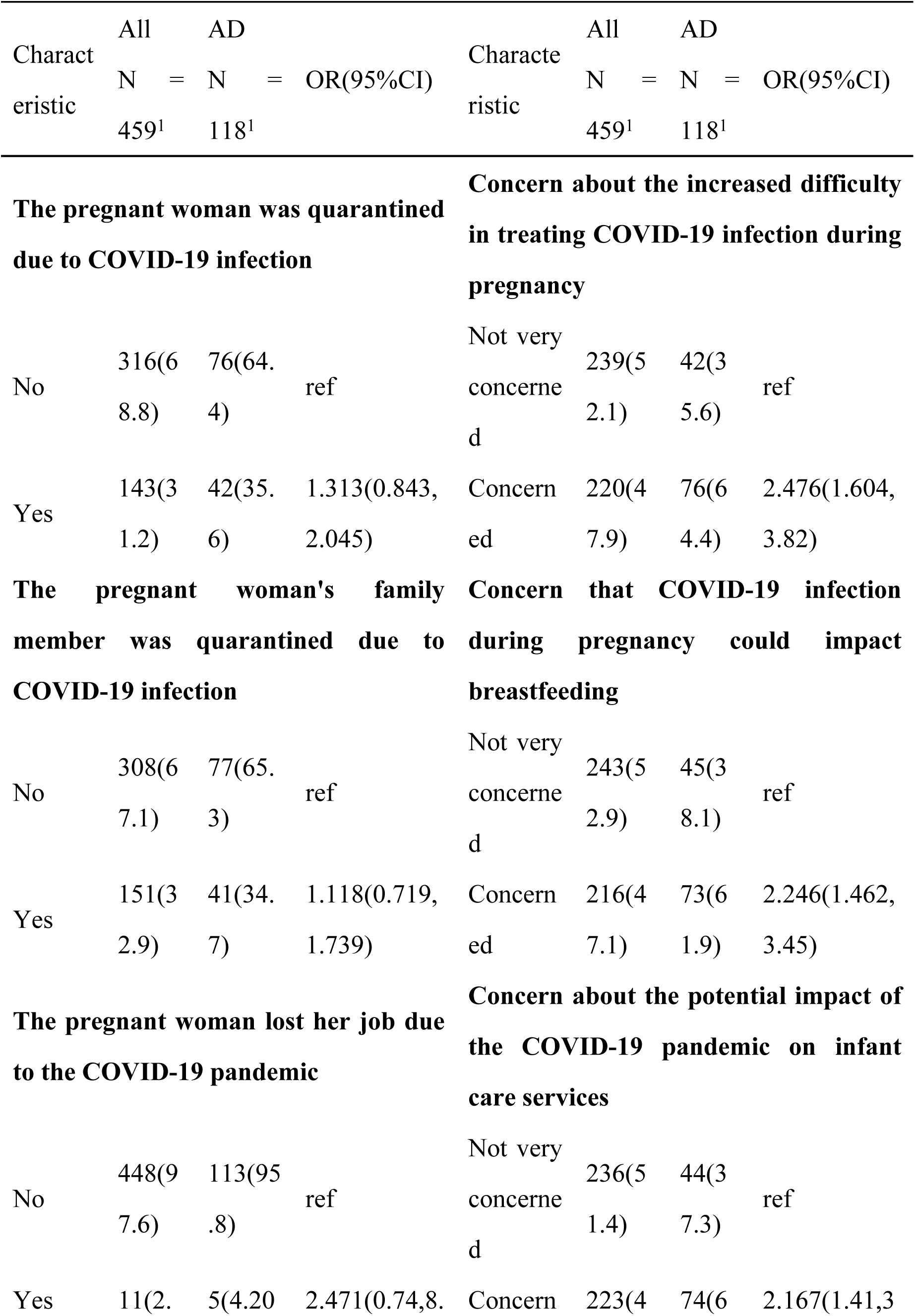
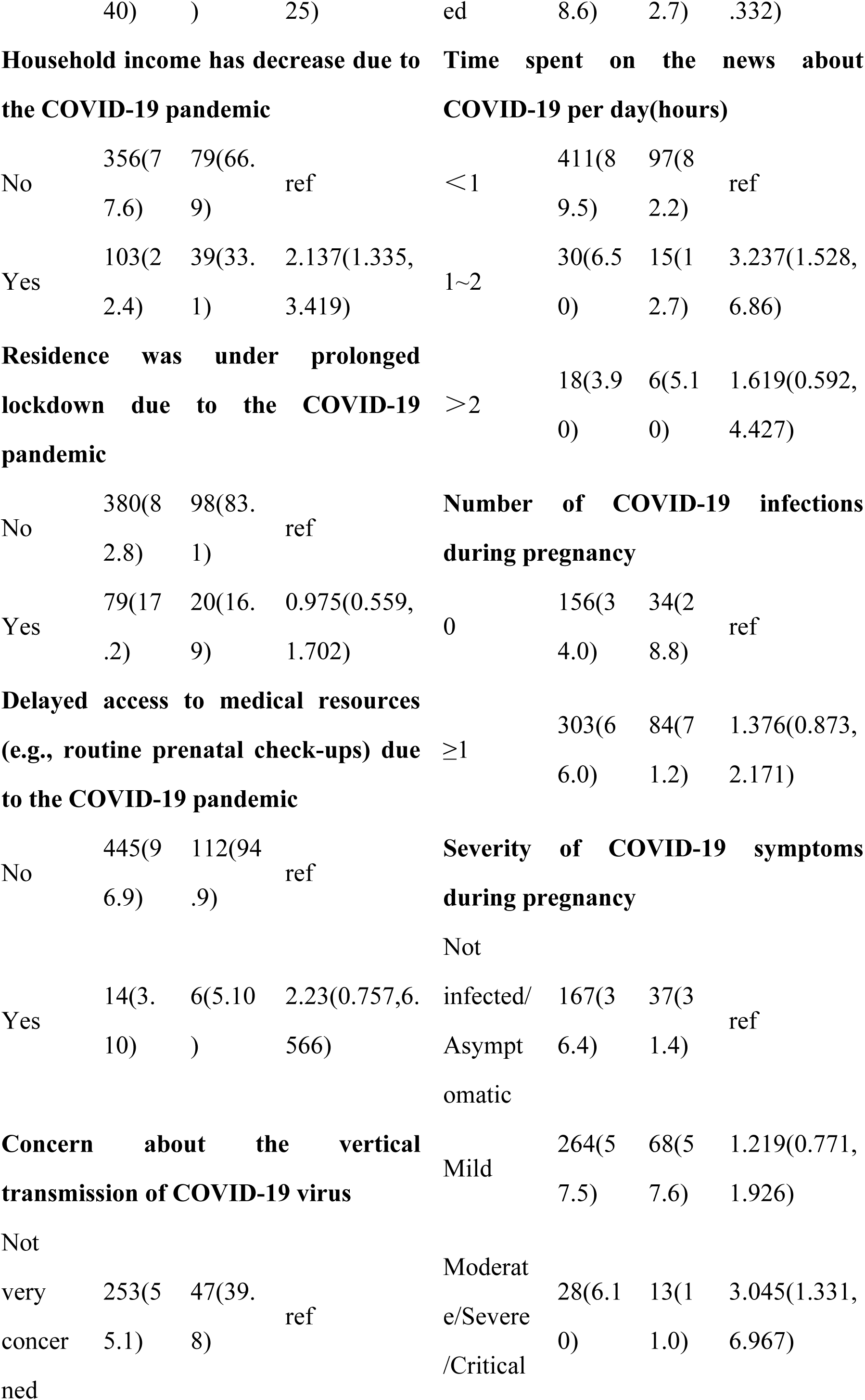
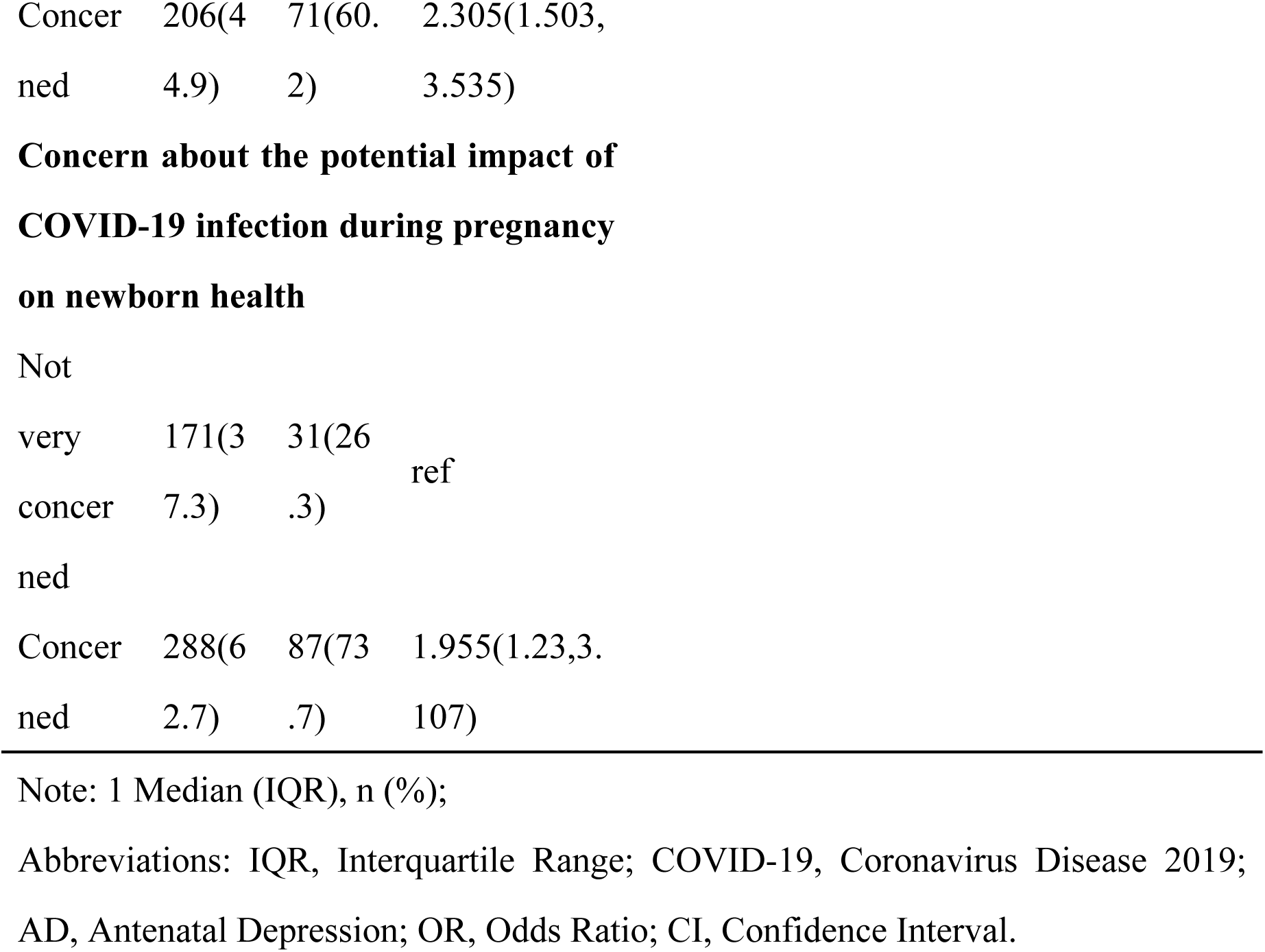
Comparative analysis of COVID-19 awareness and impact between women with AD and women with Non-AD.

In terms of pregnancy-related factors, the AD group manifested a higher prevalence of sleep disorders (PSQI ≥ 7) during pregnancy, a lower proportion living only with their husbands, and a reduced proportion with high family support, all these differences were statistically significant (*P* < 0.05). Conversely, no statistically significant differences emerged between the two groups regarding whether the pregnancy was unintended, method of conception, delivery history, presence of early pregnancy symptoms, presence of pregnancy complications, smoking during pregnancy, drinking during pregnancy, Consistency of family perception on antenatal health and care, primary decision-maker for antenatal care, proportion of pregnant women who received antenatal mental health education from professional institutions, and proportion of pregnant women who sought help from professional institutions for psychological issues (*P* > 0.05).

Regarding COVID-19-related factors, 66.0% (303/459) had contracted the COVID-19 virus at least once. Compared to the non-AD group, the AD group had a higher percentage expressing concern about the vertical transmission of COVID-19 virus, as well as the potential impact of COVID-19 infection during pregnancy on newborn health. Moreover, a larger proportion of the AD group expressed concern about the increased difficulty in treating COVID-19 infection during pregnancy, and the pandemic’s potential influence on breastfeeding as well as infant care services. The AD group also spent less time daily on the news about COVID-19, experienced a higher incidence of decreased household income due to the pandemic, and a larger proportion experienced moderate to critical symptoms post-infection. These variations were statistically significant (*P* < 0.05). However, no statistically significant differences were found between the two groups regarding factors such as personal/family members’ quarantine due to COVID-19 infection, loss of employment due to COVID-19, prolonged lockdown due to COVID-19, delayed access to medical resources due to COVID-19 (e.g., completing prenatal examinations on time), and the frequency of COVID-19 infection during pregnancy (*P* > 0.05).

### 3.3 Comparison of predictive performance of models for antenatal depression

Regarding the AD and non-AD groups as binary outcome variables, the results of the multivariate logistic regression revealed that the presence of sleep disorders (versus absence, OR=4.19, 95% CI=2.55-6.90, *P*<0.001), middle-level family support (versus high-level family support, OR=2.84, 95% CI=1.68-4.81, *P*=0.001), and experiencing moderate to severe COVID-19 symptoms during pregnancy (versus not infected/asymptomatic, OR=3.63, 95% CI=1.36-9.67, *P*=0.007) were independent risk factors for AD (results not shown).

The three significant variables identified in the multivariate analysis were utilized as independent predictive factors for establishing risk prediction models employing LR, SVM, KNN, RF, XGBoost, and GBDT machine learning algorithms. The performance of these models, in terms of precision, recall, accuracy, F1 score, AUC, sensitivity, and specificity, was summarized in Table 3. ROC curves of various models predicting AD in the training and test datasets were depicted in Figure 1A and Figure 1B, respectively. Notably, in the training dataset, the order of AUC values was RF (0.842) > GBDT (0.834) > XGBoost (0.801) > LR (0.790) > SVM (0.776) > KNN (0.749), while in the test dataset, it was RF (0.724) > XGBoost (0.713) > GBDT (0.694) > SVM (0.680) > LR (0.679) > KNN (0.599). The RF model demonstrated superior performance in both datasets, thus exhibiting the highest predictive ability, indicating its efficacy in distinguishing pregnant women in the AD group from those in the non-AD group based on the EPDS scores in this study.

**Figure 1.**
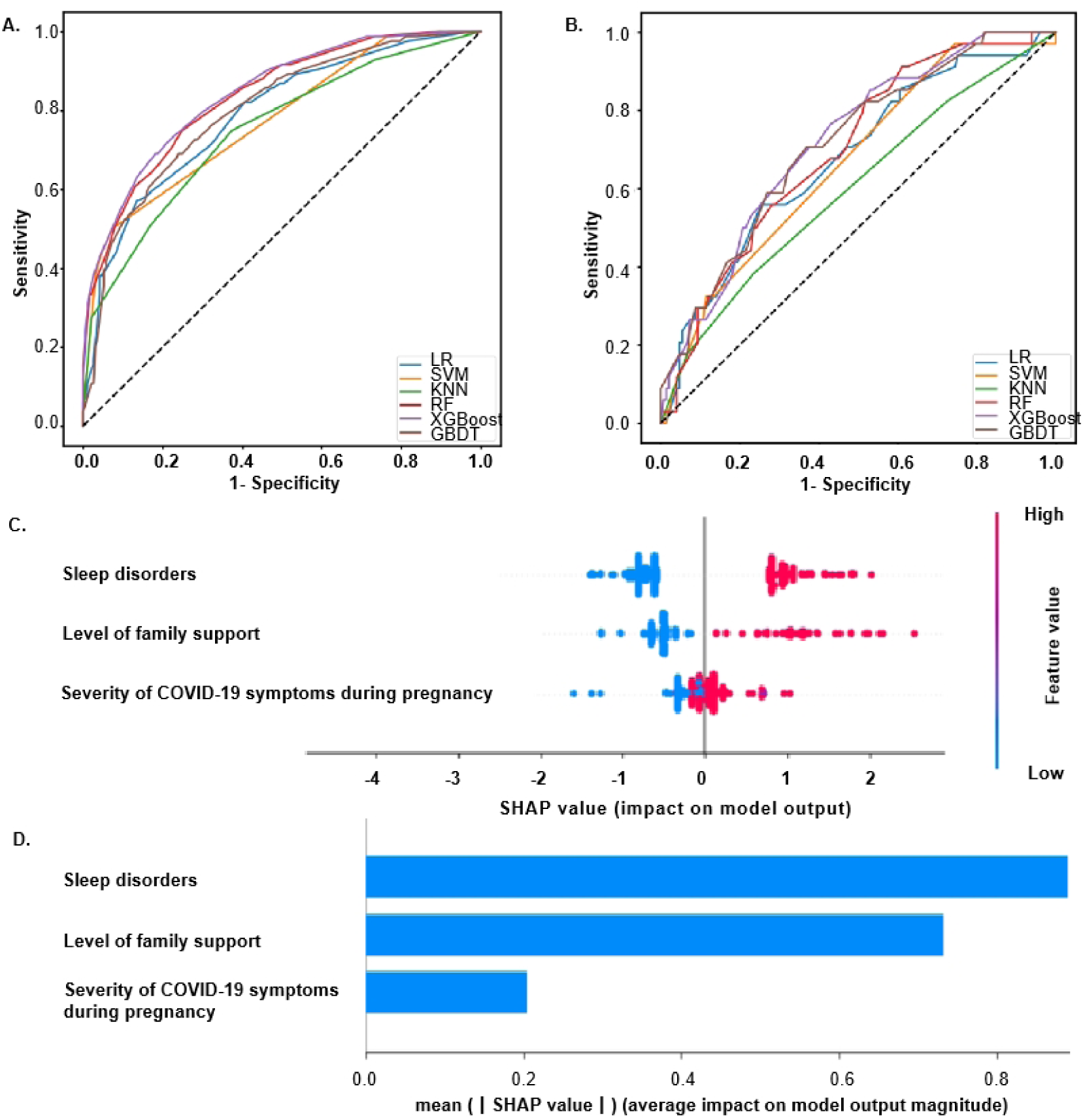
(A) ROC curves of six machine learning models for AD risk prediction in the training dataset. (B) ROC curves of six machine learning models for AD risk prediction in the test dataset. (C) ∼ (D) Relative importance of predictors based on SHAP for the RF prediction model. Abbreviations: AD, Antenatal Depression; COVID-19, Coronavirus Disease 2019; LR, Logistic Regression; SVM, Support Vector Machine; KNN, K-Nearest Neighbors; RF, Random Forest; XGBoost, eXtreme Gradient Boosting; GBDT, Gradient Boosting Decision Tree; ROC-AUC, Receiver Operating Characteristic Area Under the Curve; SHAP, Shapley’ s Additive Explanations.

**Table 3.** The comparison of predictive performance of models for AD risk.

| Indicator | Training dataset |  |  |  |  |  |
| --- | --- | --- | --- | --- | --- | --- |
|  | LR | SVM | KNN | RF | XGBoost | GBDT |
| **Precision** | 76.73% | 81.09% | 75.25% | 79.71% | 78.26% | 78.99% |
| **Recall** | 73.00% | 80.33% | 75.25% | 78.28% | 77.39% | 77.46% |
| **Accuracy** | 76.73% | 81.09% | 74.18% | 79.71% | 78.26% | 78.99% |
| **F1 Score** | 84.91% | 87.68% | 84.33% | 86.47% | 85.78% | 86.06% |
| **ROC-AUC** | 0.790 | 0.776 | 0.749 | 0.842 | 0.801 | 0.834 |
| **Sensitivity** | 38.75% | 47.50% | 16.25% | 48.80% | 41.67% | 46.43% |
| **Specificity** | 92.31% | 94.87% | 97.95% | 93.23% | 94.27% | 93.23% |
|  | Test dataset |  |  |  |  |  |
|  | LR | SVM | KNN | RF | XGBoost | GBDT |
| **Precision** | 77.60% | 75.00% | 78.26% | 78.14% | 78.14% | 77.60% |
| **Recall** | 60.83% | 60.20% | 56.55% | 60.89% | 61.59% | 61.50% |
| **Accuracy** | 77.60% | 75.00% | 78.26% | 78.14% | 78.14% | 77.60% |
| **F1 Score** | 86.56% | 84.56% | 87.65% | 87.01% | 86.93% | 86.47% |
| **ROC-AUC** | 0.679 | 0.680 | 0.599 | 0.724 | 0.713 | 0.694 |
| **Sensitivity** | 29.41% | 31.58% | 5.26% | 26.47% | 29.41% | 32.35% |
| **Specificity** | 88.59% | 86.30% | 97.26% | 89.93% | 89.26% | 87.92% |

Abbreviations: AD, Antenatal Depression; LR, Logistic Regression; SVM, Support Vector Machine; KNN, K-Nearest Neighbors; RF, Random Forest; XGBoost, eXtreme Gradient Boosting; GBDT, Gradient Boosting Decision Tree; ROC-AUC, Receiver Operating Characteristic Area Under the Curve.

### 3.4 Enhanced examination of random forest model performance

Based on the RF model, SHAP was employed to rank the importance of each variable affecting the risk of AD. The analysis revealed that “sleep disorders” had the most significant impact, followed by “level of family support” and “severity of COVID-19 symptoms during pregnancy”. (Figures 1C and 1D). Additionally, the sensitivity and specificity of the RF model were further visualized via the confusion matrix (Figures 2A and 2B).

**Figure 2.**
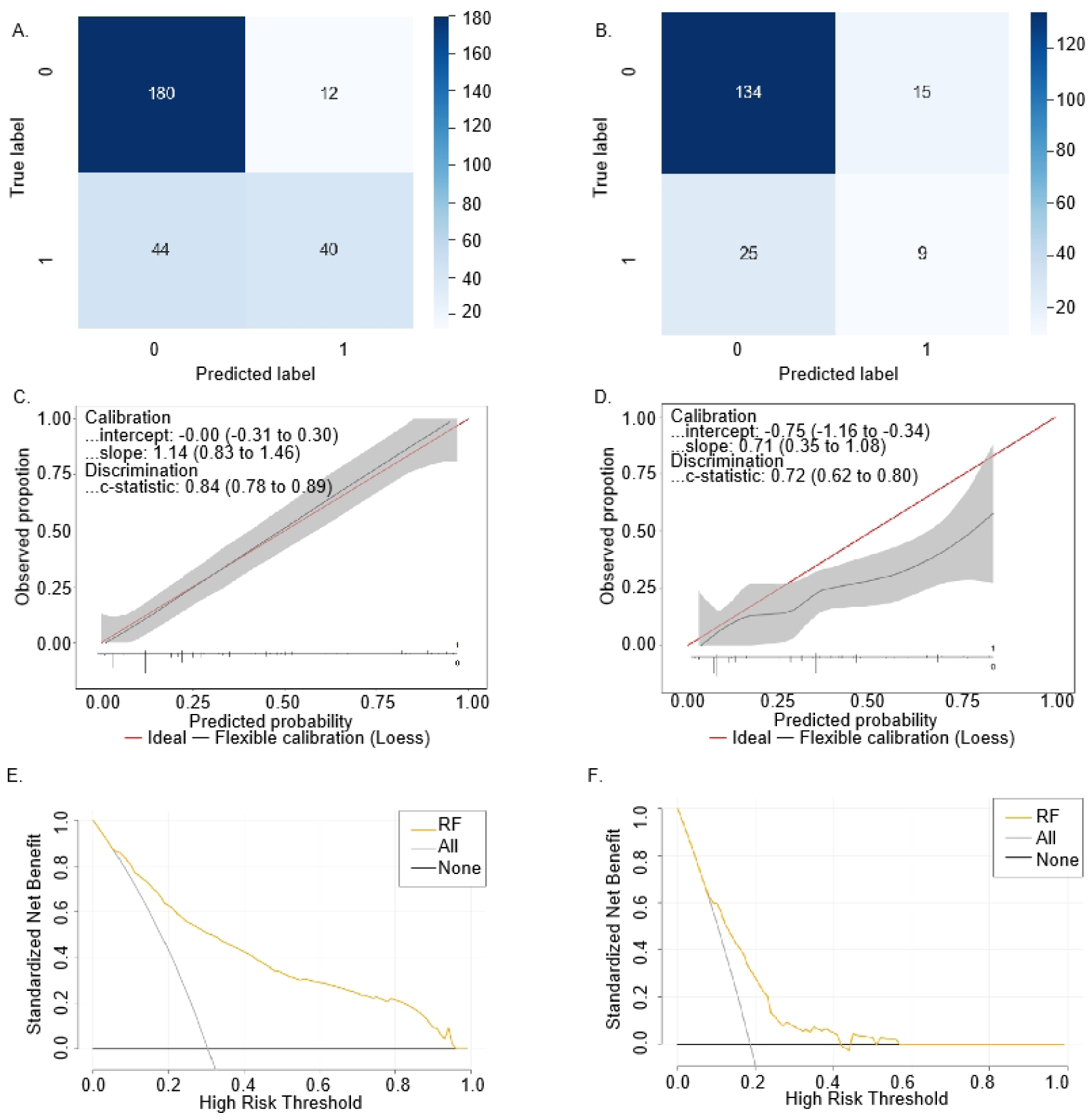
(A) CM of the RF model in the training dataset. (B) CM of the RF model in the test dataset. (C) Calibration plots of observed versus predicted AD risk for the RF model in the training dataset. (D) Calibration plots of observed versus predicted AD risk for the RF model in the test dataset. (E) DCA curve of the RF model in the training dataset. (F) DCA curve of the RF model in the test dataset. Abbreviations: CM, Confusion matrix; RF, Random Forest; AD, Antenatal Depression; COVID-19, Coronavirus Disease 2019; DCA, Decision curve analysis.

The CC demonstrated good consistency in both the training and test datasets, supported by Hosmer-Lemeshow goodness-of-fit tests of 0.797 and 0.781, respectively (*P* > 0.05). It also showed no significant statistical difference between the predicted probabilities and the observed values, with the Y-axis representing the actual occurrence status of AD and the X-axis depicting the predicted probability of AD (Figures 2C and 2D). The results of DCA for the RF model were presented in Figure 2E and Figure 2F, respectively, indicating a potential clinical benefit when utilizing the RF model to predict the risk of AD. Specifically, within certain threshold—ranging from 8% to 95% in the training dataset and 10% to 58% in the test dataset—the model demonstrates significant predictive value.

## 4. Discussion

The percentage of pregnant women testing positive for AD in this study was 25.7%, which notably exceeded the pre-pandemic domestic range of 12.5%-18.9% [7–8], yet fell within the 27.4%-39.39% range reported both domestically and internationally during the initial outbreak of pandemic [12, 15]. This suggests that the level of depression among pregnant women during the special period subsequent to the easing of pandemic lockdown was comparable to that observed during the early stage of the pandemic, thus necessitating a detailed discussion. As technology advances rapidly in data acquisition, the analysis of high-dimensional omics data has emerged as a hotspot. However, traditional statistical methods appear to be limited in this domain [16–17], while AI has shown broad prospects. ML is an essential branch of AI that is widely applied in various areas, including the development of precision medical systems, surgical assistance, and postoperative efficacy evaluation [18–19]. Although ML has become an important tool in medical practice, there are very few ML models predicting negative emotional risks in pregnant women related to the pandemic. For example, a study employed four different ML algorithms to assess the impact of the COVID-19 pandemic on the perinatal mental health of an international prospective cohort from 11 European countries. In this study, the authors set the threshold of EPDS to 13 and hypothesized that COVID-19-related factors, such as exposure to and/or a positive diagnosis of COVID-19, would be associated with an increased risk of perinatal depression [20]. Another model aimed to identify post-traumatic stress disorder (PTSD) following childbirth during the COVID-19 era, achieving a higher AUC of 0.75 compared to a traditional model with an AUC of 0.53 [21]. In China, the result of a recent AD risk prediction model based on the RF algorithm showed that antenatal anxiety, preexisting mental illness, concerns about financial costs, and family gender expectations for the fetus were risk factors for AD [22]. Another study established a binomial LR model and identified that adequate nutrition and appropriate exercise helped alleviate depressive symptoms in pregnant women during the COVID-19 pandemic [23]. However, the limited studies available primarily focused on PTSD or postpartum depression, with investigation periods concentrated in the early stages of the COVID-19 pandemic. Comparative studies using multiple ML models predominantly centered on Caucasian populations in Europe and the United States, while no similar research targeting Chinese pregnant populations. There is an urgent need for studies employing various ML models to determine the most effective approach for accurately identifying the predictors of AD among Chinese pregnant women during the special period subsequent to the easing of pandemic lockdown. Moreover, previous studies have primarily focused on developed areas, whereas the current study was simultaneously conducted in Chaoyang District in Beijing and Qiandongnan Prefecture in Guizhou Province, resulting in a notably high proportion of ethnic minorities (33.6%) among pregnant women. This offers valuable empirical evidence for formulating pertinent policies in areas with concentrated ethnic minorities

Among the six types of ML models established in this study—LR, SVM, KNN, RF, XGBoost, and GBDT—RF exhibited a generally higher AUC value compared to the other models, indicating a stronger ability to differentiate when predicting AD risk based on EPDS scores. RF was first proposed in 2001 and used decision trees as its basic units, achieving efficient classification and prediction through ensemble learning strategies [17]. By utilizing the bootstrap resampling method, RF extracts multiple subsets from the original sample set and constructs a decision tree for each subset. Then, it combines the predictions from multiple decision trees and derives the final prediction result through voting, resulting in high predictive accuracy, good tolerance to outliers and noise, and effective avoidance of overfitting. Therefore, RF is widely used in frontier fields such as data mining and bioinformatics, becoming a powerful tool for nonlinear modeling. Nonetheless, the RF algorithm is often criticized for its inherent “black box” nature, which makes it difficult to intuitively explain its internal mechanisms and hinders the interpretability of the model [17]. This limitation somewhat restricts the broader application of RF in fields like medical diagnotherapy. To enhance the interpretability of the RF model, this study intended to use the SHAP method to elucidate predictors importance. Introduced by a study in 2022, SHAP aims to elucidate the specific impact of each variable on the outcome in black-box models, thereby contributing to enhancing the model’s interpretability [24]. The findings of SHAP in this study underscored “sleep disturbances” as the most influential factor in predicting AD risk, followed by “level of family support during pregnancy” and “severity of COVID-19 symptoms during pregnancy.” These insights not only deepen our understanding of AD risk factors, but also emphasize the potential of SHAP in elucidating complex model behavior.

Meta-analysis has shown that approximately 38.2% of pregnant women experience insomnia symptoms, 15% suffer from Obstructive Sleep Apnea (OSA), and 20% are affected by Restless Leg Syndrome [25]. Numerous studies have confirmed a close association between sleep disorders and AD, indicating that sleep disorders serve as predictive risk factors for the occurrence of AD, while AD symptoms also deteriorate the sleep quality of pregnant women [25–27]. The multivariate logistic regression results of this study revealed that pregnant women with sleep disorders face a significantly increased risk of developing AD, which is 4.19 times higher than those without sleep disorders. Consistent with this conclusion, a recent meta-analysis involving 39,574 pregnant women demonstrated that compared to pregnant women with good sleep quality, those with poor sleep quality had a 3.72 times higher risk of AD [26]. Another study identified that insomnia and hypersomnia during pregnancy increase the likelihood of AD [27]. Physiological and psychological issues during pregnancy, such as hormonal changes (e.g., a sharp increase in progesterone), increased nocturnal urination frequency, physical discomfort, or emotional stress, not only exacerbate pre-existing sleep issues but can also trigger new sleep disorders [25, 27]. Additionally, sleep disorders can disrupt circadian rhythms, thereby adversely affecting the body’s immune system and inflammatory responses [28]. Pregnant women with OSA often have overweight or obesity, and a high BMI may increase AD risk through inflammatory mediators such as Glycoprotein Acetyls, Interleukin-6, and eosinophil chemotactic factor [29]. These studies provide evidence for explaining the increased risk of AD in pregnant women with sleep disorders, offering perspectives for understanding the pathogenesis of AD. Healthcare professionals and family members should pay attention to and properly address various factors that may lead to sleep disorders, in order to prevent AD.

Family factors play a pivotal role in the onset, progression, and consequences of AD in pregnant women. According to this study, pregnant women with middle level of family support during pregnancy exhibited a 2.84-fold increased risk of developing AD compared to those with high level of family support. Previous studies have reported that family cohesion and the satisfaction of pregnant women’s autonomy needs are key predictors of AD, with heightened family cohesion levels—manifesting as connected or enmeshed or types—mitigating AD incidence by bolstering pregnant women’s autonomy satisfaction [30]. The enforcement of lockdown measures during COVID-19 pandemic has increased the amount of time pregnant women spent with their families, potentially amplifying the influence of family factors on maternal mental health [31]. Prolonged lockdown may expose pregnant women to challenges such as social isolation, disrupted daily routines, and inability to access prenatal care and medical services, thereby elevating their psychological stress. Moreover, adjustments to pandemic lockdown policies bring changes, such as purchasing pandemic prevention supplies, seeking pandemic-related consultations and treatments, which significantly test the cooperation of family members. High level of family support can effectively alleviate these pressures for pregnant women. Therefore, in responding to major emergencies and their policy adjustments, it is crucial to consider the potential impact of family factors on the mental health of pregnant mothers. Effective interventions and support strategies should be established to strengthen family unity.

Compared to uninfected or asymptomatic pregnant women, those with mild COVID-19 symptoms in this study had a 1.32 times higher risk of AD (*P*=0.207), while those with moderate, severe or critical symptoms had a 3.63 times higher risk (*P*=0.007). Except that more severe symptoms of COVID-19 infection psychologically exacerbate pregnant women’s concerns about the virus’s impact on fetus health, it also suggests that COVID-19 infection may elevate the risk of AD by directly affecting maternal health. However, the underlying mechanism remains unclear and may be linked to pathological changes in the cerebral cortex [32–35]. Prior to the pandemic, a study used voxel-based whole-brain analysis with resting-state functional magnetic resonance imaging to investigate differences in fractional amplitude of low-frequency fluctuation (fALFF) between AD patients and healthy pregnant women. They discovered that patients with AD showed elevated fALFF in specific brain regions, including the left medial prefrontal cortex, dorsolateral prefrontal cortex, and anterior cingulate cortex. Conversely, reduced fALFF was observed in the bilateral orbitofrontal cortex, the right insula, a cluster encompassing the right ventral temporal cortex (VTC), and the para-hippocampus (PHP). Notably, the EPDS scores of AD patients exhibited a negative correlation with fALFF values in the right VTC and PHP [32]. During the early pandemic, a study examined single-nucleus transcriptomes from the medial frontal cortex of 8 COVID-19 patients revealed widespread cellular perturbations. Specifically, this study observed the infiltration of peripheral T cells into the brain tissue, and distinct microglia and astrocyte subpopulations associated with COVID-19 exhibited similarities to pathological states seen in neurodegenerative diseases. Notably, synaptic signaling in upper-layer excitatory neurons, which are evolutionarily expanded in humans and crucial for cognitive function, wass preferentially impacted by COVID-19. These COVID-19-related perturbations overlap with those seen in chronic brain disorders and are associated with genetic variants linked to depression [33]. A case-control study employed positron emission tomography to measure the translocator protein distribution volume (TSPO VT) in patients with persistent depressive and cognitive symptoms after mild to moderate COVID-19 infection, founding elevated TSPO VT, indicating increased gliosis, particularly in the ventral striatum and dorsal putamen when compared to healthy controls [34]. Furthermore, a recent study evaluated the neuropsychological profile of patients suffering from asymptomatic to moderate acute COVID-19. Its findings revealed multidomain cognitive impairment, including learning/memory and executive functions, complex attention deficits, language difficulties, and perceptual-motor dysfunction, documenting cognitive deficits as a component of the post-COVID-19 syndrome. These findings provide insight into the potential psychopathology of how COVID-19 infection might damage the brain structure of pregnant women, leading to an increased risk of AD [35].

Despite numerous innovations, this study has the following limitations: Firstly, due to the cross-sectional study design, causal relationships between influencing factors and AD cannot be directly determined. Secondly, data from scales such as PSQI may be subject to recall bias, and the subjectivity of the EPDS scale may lead to biased assessment results, potentially affecting the accuracy and reliability of the data. Thirdly, the EPDS threshold set in this study was based on a pre-pandemic study conducted on Chinese pregnant women. While supported by literature, its applicability to pregnant women from diverse cultural and social backgrounds may vary. Fourthly, given the intricacies of the RF algorithm as a complex ensemble learning technique, although this study provides SHAP feature importance assessment, calibration curves, and clinical decision curves to enhance the predictive power of the RF model, its interpretability and generalizability in clinical applications may be limited. Therefore, caution is warranted when interpreting and applying the predictive results.

## 5. Conclusion

In summary, sleep disorders, middle-level family support, and experiencing moderate to severe COVID-19 symptoms during pregnancy emerged as independent risk factors for AD following the easing of pandemic lockdown. Implementing comprehensive strategies to effectively address these factors holds significant meaning. Among the various ML models established for AD risk prediction using these three predictors, the RF model exhibited superior effectiveness in distinguishing whether pregnant women have AD, thereby providing a targeted tool for screening AD during the post-lockdown period. This can serve as a valuable reference for policy adjustments during public health emergencies, aiding in alleviating AD-related medical burdens and saving medical costs. To conclude, the integration of ML into AD risk assessment shows promise for enhancing early detection and intervention strategies. Additionally, SHAP provides a viable visual interpretation of the impact of these variables on AD occurrence, elucidating the role of individual features in prediction outcomes.

## Author contributions

**Yijin Pan:** Conceptualization, Writing - original draft, Formal analysis. **Hai Lin:** Methodology, Data curation, Formal analysis. **TAKAE HIRONO:** Data curation. **Yifan Yang:** Data curation. **Yang Liu:** Data curation. **Yinqiang Zhang:** Conceptualization, Writing - review and editing, Supervision.

## Data availability

The datasets generated during and analyzed during the current study are available from the corresponding author on reasonable request.

## Acknowledgements

We thank Xiaoyu Pan, M.D. from the Department of Obstetrics and Gynecology, China-Japan Friendship Hospital, Beijing, China, for her valuable advice and critical support throughout the study.

